# Digenic inheritance of mutations in *SPG7* and *AFG3L2* causes motor neuron and cerebellar disorders

**DOI:** 10.1101/2025.07.05.24312261

**Authors:** Mehrdad A Estiar, Eric Yu, Parizad Varghaei, Jay P. Ross, Setareh Ashtiani, Andrew N. Bayne, Giulia Coarelli, Dagmar Timmann, Thomas Klockgether, Danique Beijer, David Mengel, Marie Coutelier, Project MinE ALS Sequencing Consortium, Patrick A. Dion, Oksana Suchowersky, Claire Ewenczyk, Cyril Goizet, Giovanni Stevanin, Philip Van Damme, Ammar Al-Chalabi, Stephan Zuchner, Matthis Synofzik, Jan H. Veldink, Jean-Francois Trempe, Alexandra Durr, Guy A. Rouleau, Ziv Gan-Or

## Abstract

**Background:** Biallelic *SPG7* mutations cause one of the most common forms of hereditary spastic paraplegia (HSP). Several reports have suggested that heterozygous *SPG7* variants may also play a role in HSP, but also in amyotrophic lateral sclerosis (ALS). However, it remains controversial whether heterozygous *SPG7* mutations are pathogenic on their own, or if other mechanisms are at play. We recently provided evidence for non-Mendelian inheritance in spastic paraplegia 7 (SPG7), as heterozygous carriers of *SPG7* mutations often also carried mutations in other disease-related genes, more frequently than expected by chance. *AFG3L2* encodes for the AFG3 Like Matrix AAA Peptidase Subunit 2 protein, which creates a proteolytic complex with SPG7-encoded protein paraplegin. In this study, we aimed to examine whether digenic heterozygous mutations in *SPG7* and *AFG3L2* can lead to a spectrum of neurodegenerative disorders.

**Methods:** We first analyzed genome and exome sequencing data of 7,515 unrelated individuals including 5,108 motor neuron disorder (MND) and ataxia patients and 2,407 controls. We next analyzed an additional 18,748 exome data from rare disease cohorts to further examine the occurrence of variants in *SPG7* and *AFG3L2*.

**Results:** Among the first 5,108 MND and ataxia patients, we identified a total of 10 patients, 8 of whom were unrelated, carried potentially pathogenic variants in both *SPG7* and *AFG3L2*, in contrast to none in 2,407 unrelated controls. Further analysis of the 18,748 additional patients with rare disease, as well as a comprehensive literature review, identified 7 more patients, 6 of whom were unrelated, had digenic mutations in *SPG7* and *AFG3L2*. In the two families we identified, digenic mutations in *SPG7* and *AFG3L2* perfectly segregated with the disease. The 17 patients reported here exhibited predominant signs of motor neuron and cerebellar involvement.

**Conclusions:** Our findings demonstrate that digenic inheritance of concurrent heterozygous mutations in *SPG7* and *AFG3L2* may cause motor neuron and cerebellar disorders. Screening of the entire *SPG7* and *AFG3L2* genes in genetically undiagnosed cases of MND and spastic ataxia may help to increase the diagnostic yield.

## Background

*SPG7* is the first identified gene in autosomal recessive hereditary spastic paraplegia (HSP), with biallelic *SPG7* mutations found in 62% of autosomal recessive HSP patients (1). *SPG7* encodes paraplegin, a mitochondrial matrix protease localized in the inner membrane of mitochondria (2), where it assembles with homologous AFG3L2 (ATPase family gene 3-like 2) subunits and forms a hetero-oligomeric proteolytic complex (SPG7-AFG3L2) (3, 4). This complex is crucial for the normal function of mitochondria and its impairment results in mitochondrial dysfunction (3, 5). SPG7 can only assemble with AFG3L3 to form this complex, whereas homo-oligomeric complexes of AFG3L2 (AFG3L2-AFG3L2) also exist.

Biallelic *SPG7* variants have been reported in multiple neurological disorders in addition to spastic paraplegia 7 (SPG7; OMIM 607259), including cerebellar and spastic ataxia (6–11), parkinsonism (12, 13), isolated optic atrophy (14), progressive external ophthalmoplegia (15), limb dystonia (16, 17), early-onset optic neuropathy (18), and primary progressive multiple sclerosis (19). Several reports suggested that heterozygous *SPG7* variants may also be pathogenic in neurological disorders, including HSP (14, 20), primary lateral sclerosis (PLS) (21, 22), and amyotrophic lateral sclerosis (ALS) (23, 24).

While heterozygous *SPG7* mutations have also been suggested to cause disease, we showed in a recent study that many of those who carried a heterozygous *SPG7* variant also carried other variants in HSP-related genes or in genes that may interact with SPG7 (25). This observation may suggest that SPG7 could be involved in digenic inherited neurological disorders rather than a monogenic effect of heterozygous *SPG7* variants. One of the HSP patients that we identified carried heterozygous variants in both *SPG7* and *AFG3L2* (25). Interestingly, a previous case report described a patient with early-onset optic atrophy, spastic ataxia, and parkinsonism who also had digenic heterozygous mutations in *SPG7* and *AFG3L2*, yet no additional patients with mutations in both genes have been reported. The case for digenic inheritance is further strengthened by different animal models with dysfunction of both proteins showing early-onset axonal degeneration, prominent cerebellar degeneration with loss of Purkinje cells and parallel fibers, reactive astrogliosis and defective mitochondria (26, 27).

In the current study, we aimed to examine whether digenic heterozygous mutations in *SPG7* and *AFG3L2* may indeed cause a spectrum of diseases in which the corticospinal and spinocerebellar tracts are affected, by analyzing genetic data from 5,108 MND and ataxia patients and 2,407 controls, followed by additional analysis of 18,748 individuals with rare diseases. We identified a total of 17 individuals with concurrent heterozygous mutations in *SPG7* and *AFG3L2* (14 of which were unrelated), and none in controls.

## Methods

### Participants

We used 7,515 next-generation sequencing (NGS) datasets (Supplementary Table 1) including 6,183 genome and 1,332 exome sequencing data. It included 4,564 unrelated ALS patients and 2,407 unrelated control individuals. The recruitment of 4,356 ALS patients and 1,827 age- and sex-matched controls were performed through the Project MinE cohort and the participants, including both cases and controls, were primarily of European ancestry (28). An additional 208 ALS patients were recruited across Québec, Canada. The exome sequencing data of 291 index HSP patients and 580 unrelated controls were already published, and details can be found in the original papers (29, 30). We also analyzed exome sequencing data from 253 ataxia patients in France, recruited as part of the SPATAX cohort. In addition to these datasets, we analyzed 12,407 NGS datasets of rare diseases from the Genesis cohort (31) as well as data from 1,341 patients with neurodegenerative disorders from Dr. Synofzik’s lab in Germany and 5,000 patients with neurodegenerative disorders seen by various neurologists throughout Germany, to find potential concurrent heterozygous variants in *SPG7* and *AFG3L2*. We performed an extensive literature search with PubMed using the keywords “SPG7” and “AFG3L2”. We examined 31 studies for co-occurrent *SPG7* and *AFG3L2* variants carriers.

### Genetic analysis

DNA was purified from peripheral blood according to standard procedures. Details on genome and exome sequencing were previously described (28, 29). For genome sequencing data, variant calls with <15X depth of coverage and a genotype quality of <97 were excluded. For exome sequencing data, variant calls with <30X depth of coverage, a genotype quality <97 and genotyping frequency < 25% were excluded from the analysis. We did relatedness tests using KING (32) and GCTA (33), separately. GCTA detected more related samples (3^rd^ degree or closer), along with all the detected individuals by KING and then we removed 15 genome sequencing data including 10 cases and 5 controls.

The initial selection of variants was based on exonic (except for synonymous) and splice-site variants in *SPG7* (OMIM 602783, NM_003119.3, GRCh37/hg19) and *AFG3L2* (OMIM 604581, NM_006796, GRCh37/hg19) with allele frequency <0.01 in gnomAD v2.1.1. Then, the variants were interpreted using VarSome (34) and according to the American College of Medical Genetics and Genomics (ACMG) and the Association for Molecular Pathology guidelines (35). The variants were classified as either benign (B), likely benign (LB), uncertain significance (VUS), likely pathogenic (LP), or pathogenic (P). We excluded variants that were classified as B/LB. The splice-site variants with VUS ±3 were also excluded and novel deletions/insertions and frameshift variants were considered as likely pathogenic.

The Manta tool was applied to call structural variants (SVs) from genome sequencing data. Multiplex ligation-dependent probe amplification (MLPA) was performed using the MLPA SALSA kit (MRC-Holland; Probemix P213 HSP mix-2) on a subset of samples that went through exome sequencing to discover copy number variations (CNVs). In order to predict the presence of important domains and sites of the corresponding protein, we used InterPro (36). A *P*-value less than 0.05 was considered statistically significant for all results using binomial test, Pearson chi-square or Fisher’s Exact test.

### Structural visualization and mutational analysis

AFG3L2-SPG7 heterohexamer models were generated using the AlphaFold3 server (37). Briefly, full length AFG3L2-SPG7 and matrix domain AFG3L2 (a.a. 272-797)-SPG7 (a.a. 279-795) heterohexamers were generated by specifying a 3:3 stoichiometry with default AlphaFold3 parameters. The resulting complex was relaxed using AMBER with 2000 iterations, a tolerance of 2.39 kcal/mol and a stiffness of 10 kcal/mol Å (37). To estimate the locations of nucleotide binding sites and metal binding, one chain of each AFG3L2 and SPG7 matrix domains were inputted to the AlphaFill database (38) and the highest confidence predictions containing Mg^2+^, ADP, and Zn^2+^ co-factors were downloaded as individual PDB files. The positioning of these co-factors was verified by comparison to the cryo-EM structure of the substrate-bound AFG3L2 homohexamer (PDB: 6NYY) (39). These individual AFG3L2/SPG7 chains from AlphaFill were then re-aligned onto the AFG3L2-SPG7 hexamer model in PyMOL v2.5.4 to position the co-factors within the entire assembly. All mutations and residue interfaces were visualized using PyMOL and annotated in Adobe Photoshop and Illustrator. To assess clashes induced by patient mutations, mutagenesis was performed in PyMOL and all rotamers were manually inspected. To support this manual mutagenesis approach, the stabilizing or destabilizing effects of individual missense mutations were also probed using DynaMut2 (40) on the unrelaxed AFG3L2-SPG7 heterohexamer model. The predicted changes in Gibbs Free Energy (ΔΔG) from DynaMut2 were manually annotated onto the structural visualization.

## Results

### Concurrent *SPG7* and *AFG3L2* mutations

We analyzed 7,515 exome and genome sequencing datasets of 4,564 ALS, 291 HSP, and 253 ataxic patients, and 2,407 control individuals to find rare P/LP (<1%) *SPG7* and *AFG3L2* variants. We found 8 index patients with co-occurring mutations vs. none in controls (Table 1). One of the index cases is an affected mother with two affected children, all of whom carry both variants, demonstrating complete segregation (Figure 1). To further examine the spectrum of potentially pathogenic concurrent *SPG7* and *AFG3L2* variants, we embarked on a larger effort to identify additional patients. We further analyzed the exome sequencing data from 12,407 individuals with rare diseases in the Genesis cohort as well as 6,341 individuals with neurodegenerative disease (NDD). We also searched for publications reporting cases with *SPG7* and/or *AFG3L2* variants. We identified six additional patients with concurrent variants. Consequently, a total of 14 index patients with co-occurring *SPG7* and *AFG3L2* variants were discovered, all presenting symptoms of cerebellar disorders. These include two ALS patients with concurrent SNVs in *SPG7* and *AFG3L2*, one ALS patient with *SPG7* SNV and heterozygous large insertion in *AFG3L2*, and one ALS patient with pathogenic nonsense *AFG3L2* SNV who harbored a heterozygous *SPG7* CNV (Table 1). In addition, five patients from the ataxia cohort in France, four from various NDD patients from Germany, one from the HSP cohort in Canada, one from Genesis, and two from other published cohorts (27, 41) carried digenic SNVs in *SPG7* and *AFG3L2.* Except for one of the *AFG3L2* SNVs, all other SNVs are either nonsense or occurred in the Peptidase M41 domain of AFG3L2 (IPR000642). The most common mutations in concurrent patients were *SPG7*:p.(Ala510Val) and *AFG3L2*:p.(Arg702Gln). In Families 5 and 9, with a total of 5 patients, digenic *SPG7* and *AFG3L2* variants co-segregated with the disease (Figure 1). In contrast, individuals carrying only one of the variants remained asymptomatic.

**Table 1.**
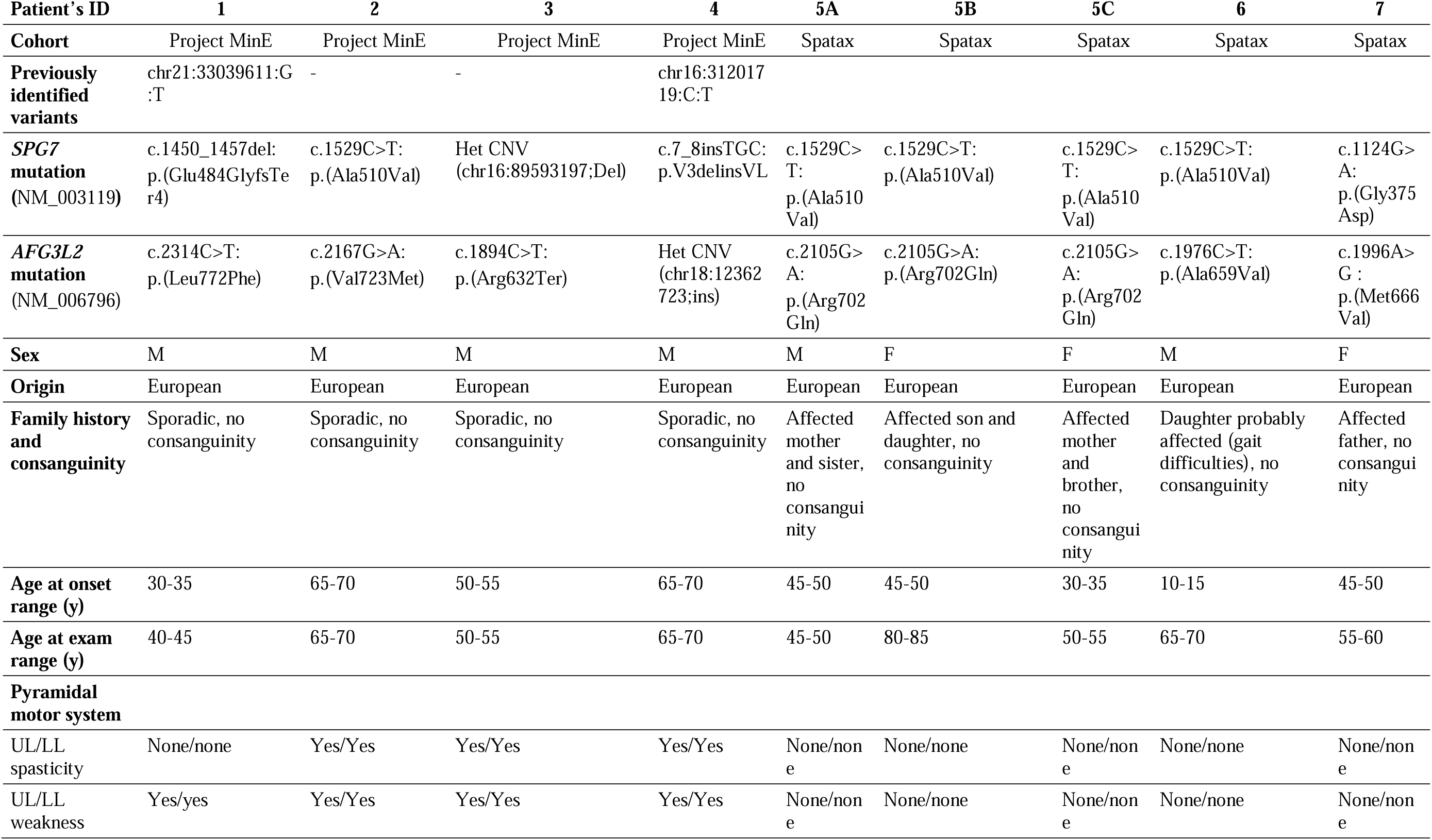
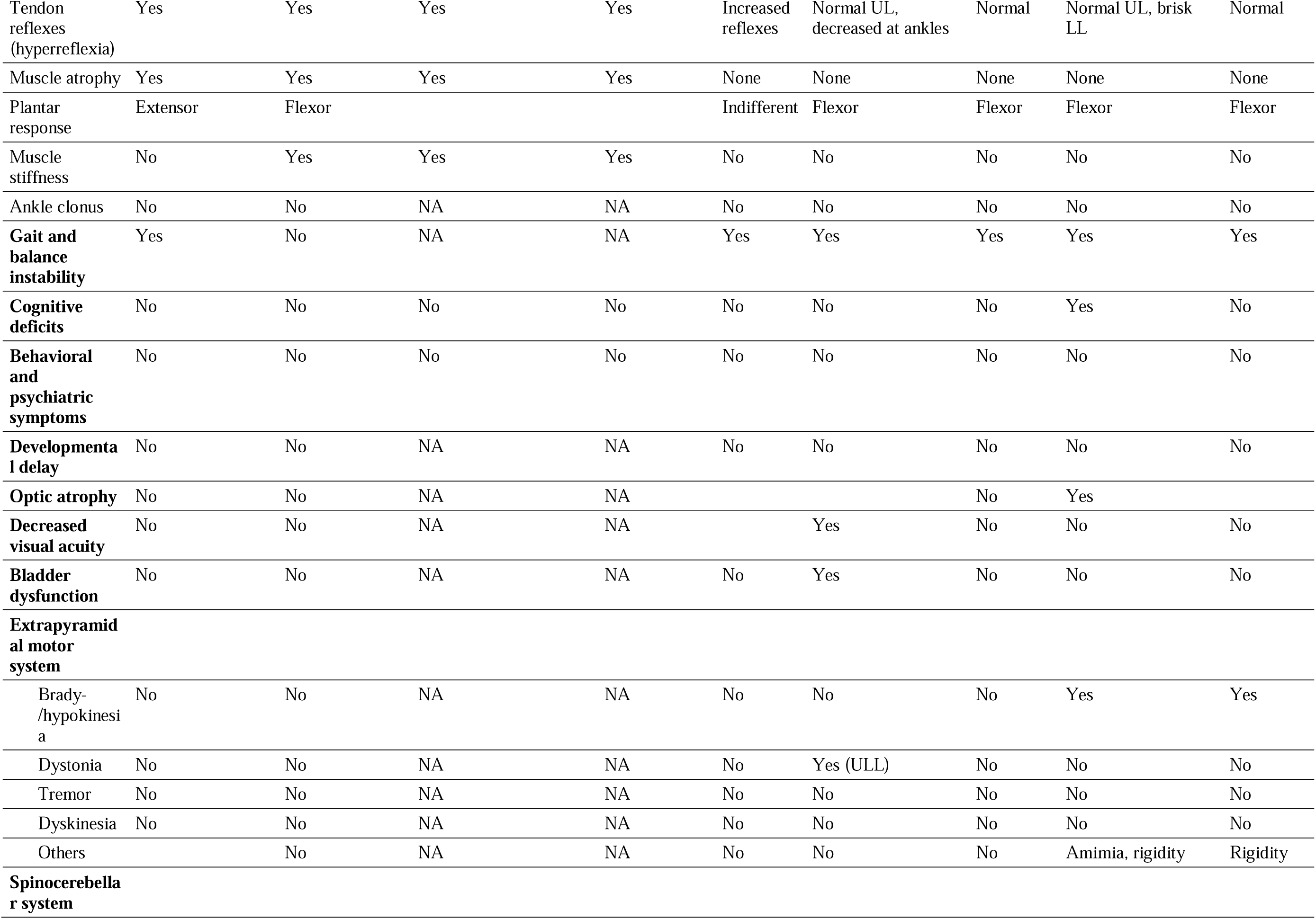
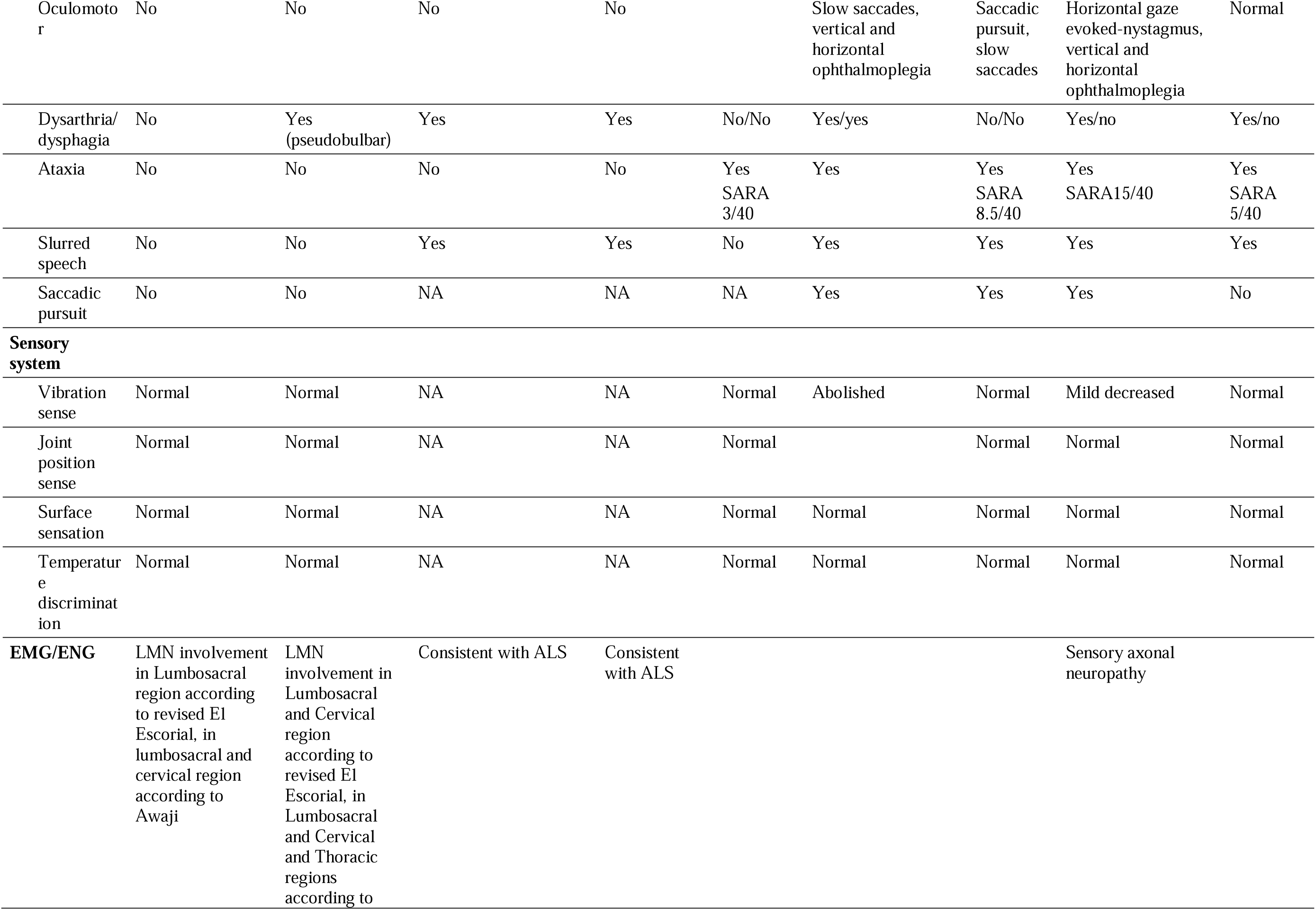
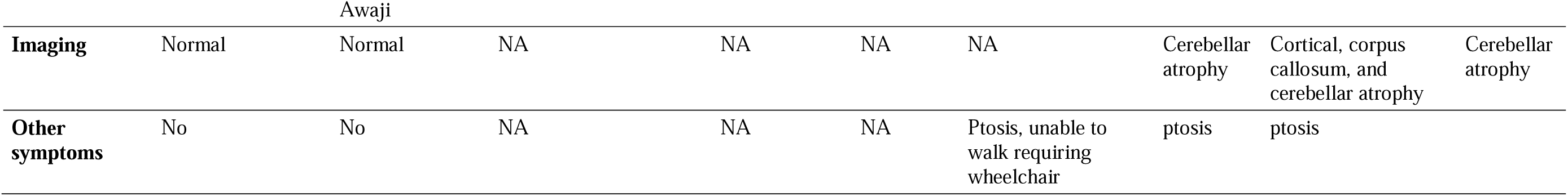
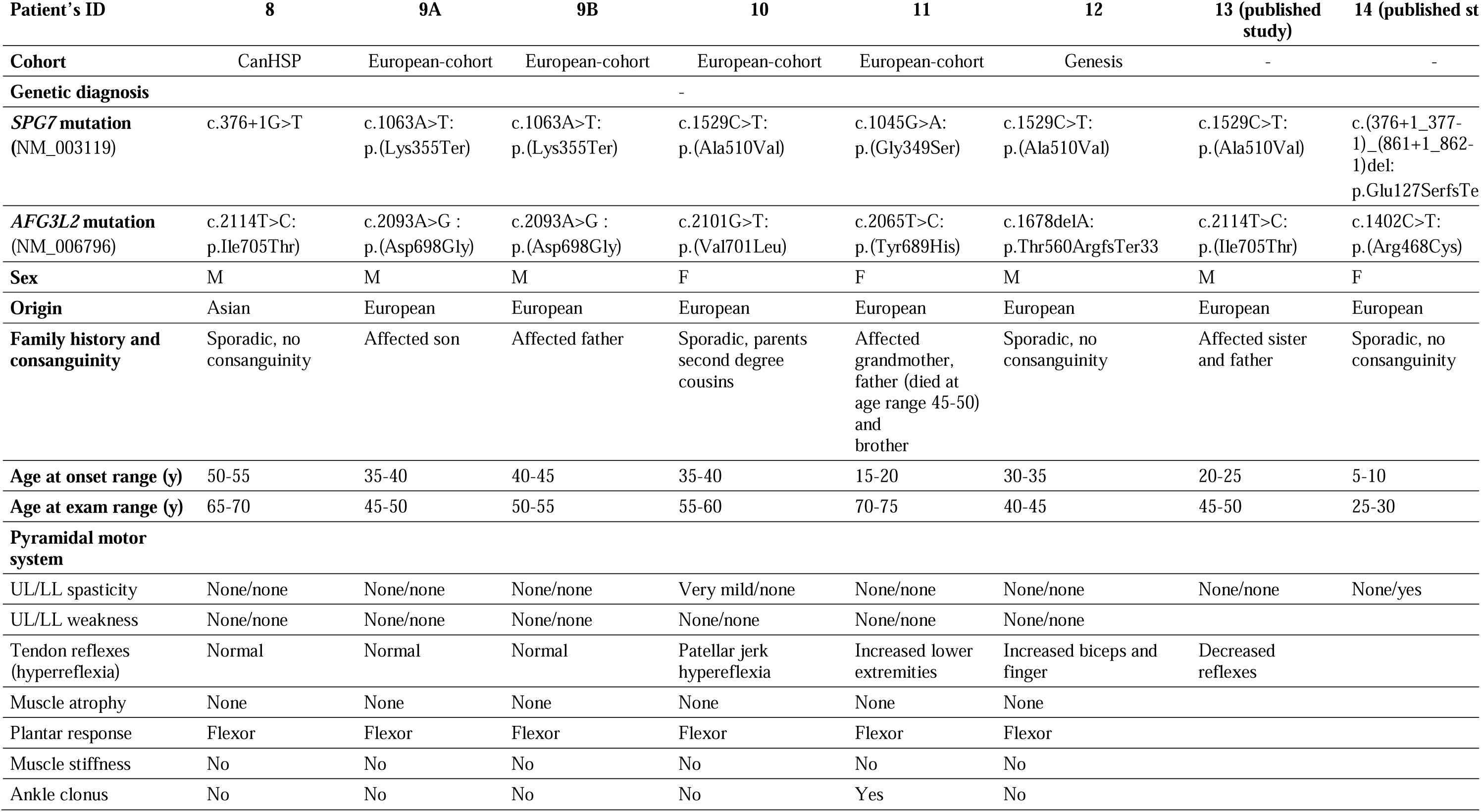
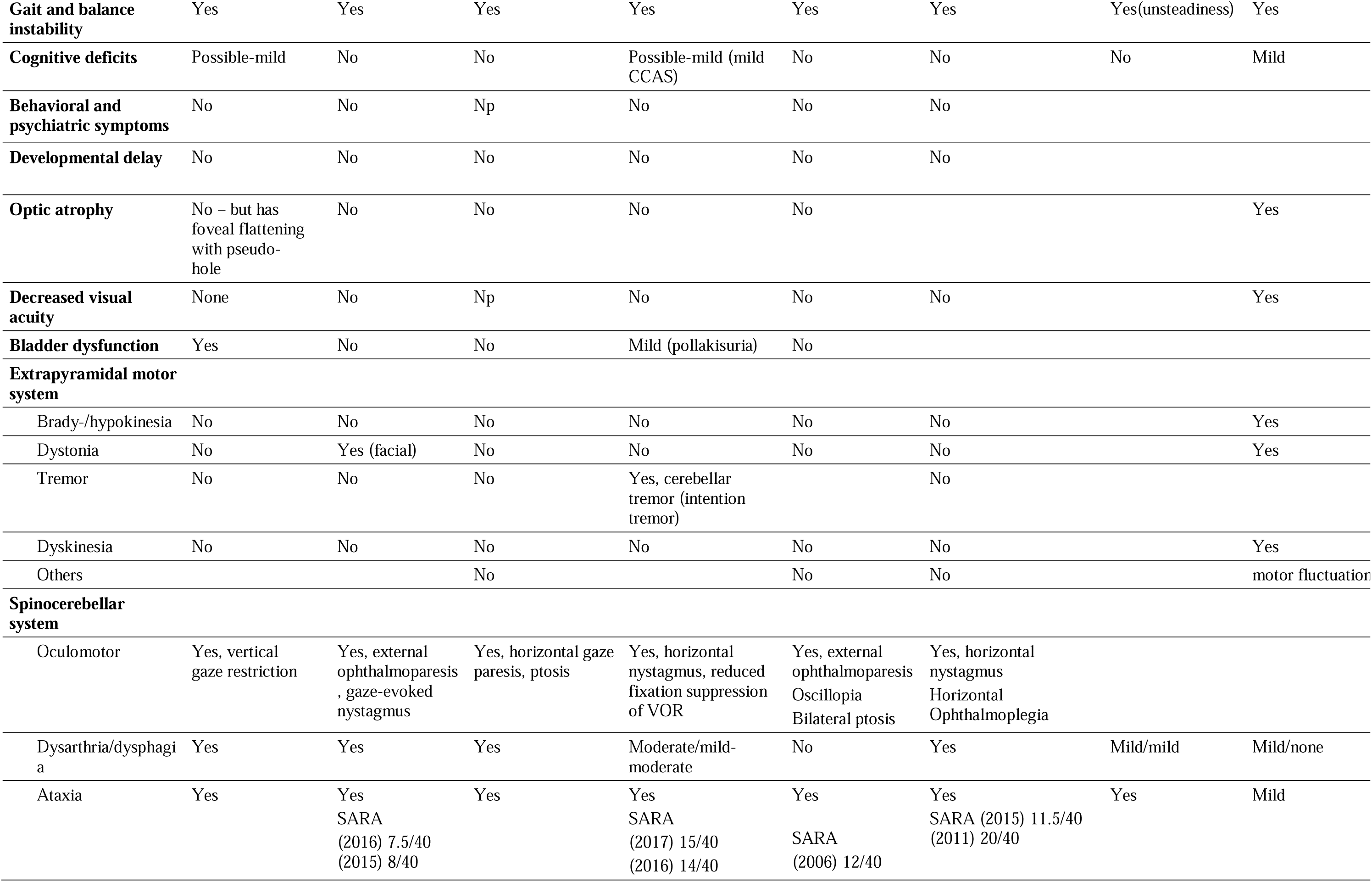
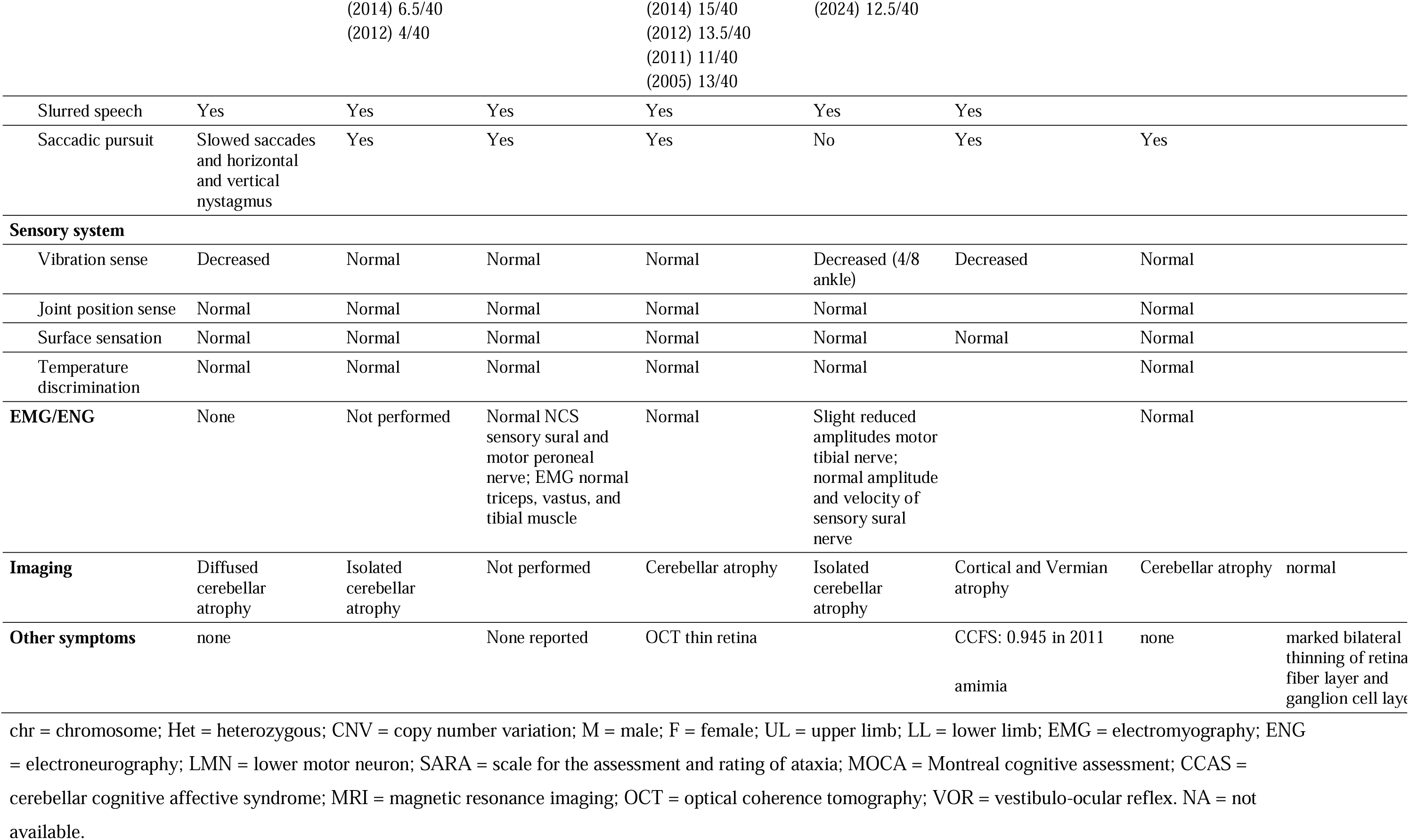
Genetic and clinical characteristics of patients with digenic mutations in *SPG7* and *AFG3L2*.

**Figure 1.**
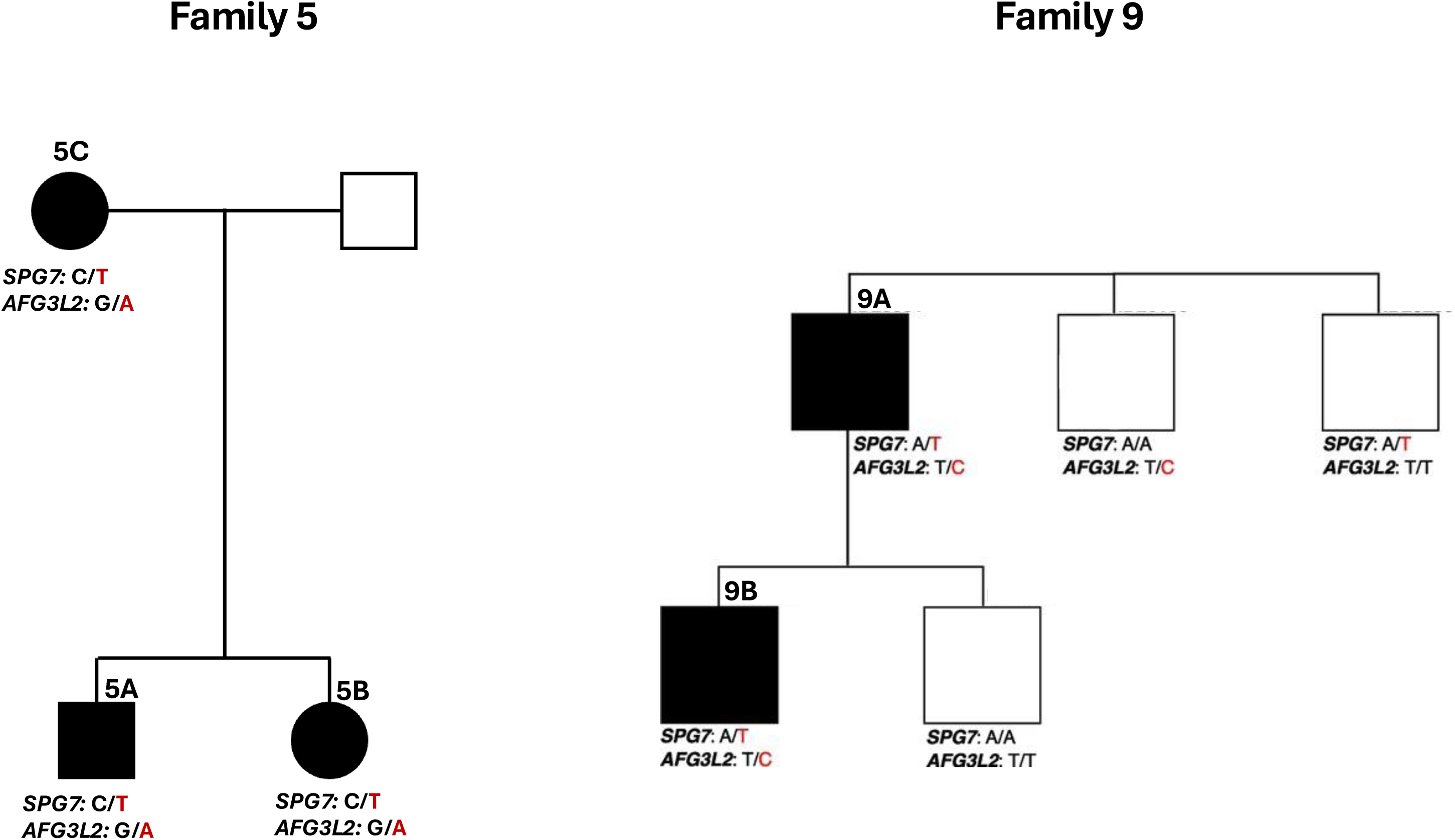
Pedigree of non-sporadic patients with available segregation data, demonstrating full segregation of the *SPG7* and *AFG3L2* variants.

### Clinical characteristics of the digenic patients

Table 1 details the clinical characteristics of all patients. The average age at onset is 38.7 years, ranging from 5 - 67, with gait difficulties as the main presenting symptom. Except for one patient, none of the concurrent patients come from consanguineous families, less than half were isolated cases (8/17), and the remaining with multigeneration cases. All but one, who was of Chinese ancestry, were of European origin, and there were more men (11/17). The clinical presentation and progression were highly variable between patients. Cognition was not affected in most patients and mildly affected in some, and no developmental delay has been reported in any of the patients. The sensory system was unaffected in most patients, but spinocerebellar signs were reported in most patients, especially oculomotor, and imaging demonstrated cerebellar atrophy in 8/12 patients in which MRI was performed. Bladder function was normal in most patients.

### Frequency of non-digenic *SPG7* and *AFG3L2* variants in ALS patients

We did not find overrepresentation of P/LP *SPG7* or *AFG3L2* alleles separately in ALS patients vs. controls (*SPG7*: 122/9128 vs. 54/4814; *P* = 0.298; *AFG3L2*: 43/9128 vs. 15/4814; *P* = 0.212; Supplementary Tables 2-3). In both cases and controls, the most common P/LP *SPG7* and *AFG3L2* alleles were p.(Ala510Val) and p.(Leu772Phe) respectively, which were not significantly different between our ALS patients and controls (*SPG7*:p.Ala510Val: 54/9128, 0.59% vs. 27/4814, 0.56 *P* = 0.823; *AFG3L2*:p.Leu772Phe: 29/9128 vs. 11/4814, *P* = 0.348). After excluding the common alleles, the number of P/LP alleles in both genes were still not different between groups (*SPG7*: 68/9128 vs. 27/4814; *P* = 0.208; *AFG3L2*: 14/9128 vs. 4/4814; *P* = 0.271; Supplementary Tables 2-3). Similarly, the rare variant burden test in *SPG7* and *AFG3L2* using the Project MinE data browser (http://databrowser.projectmine.com/) also did not show a significant association (*SPG7 P* = 0.324; *AFG3L2 P* = 0.937; Supplementary Figure 1).

To assess the co-occurrence of rare variants in *SPG7* and *AFG3L2*, a binomial test was performed (42). The analysis compared the occurrence of variants in 5,464 ALS patients and 2,407 controls. In the ALS group, 118 individuals carried rare *SPG7* variants and 39 carried rare *AFG3L2* variants, while in the control group, 54 individuals carried *SPG7* variants and 15 carried *AFG3L2* variants. The observed co-occurrence of these variants in ALS patients was significantly lower than expected under the null hypothesis of independent occurrence, with a p-value of 2.2 × 10 ¹, suggesting a significant association between the variants in ALS patients compared to controls.

### Structural analysis of AFG3L2/SPG7 hexamers and digenic mutations

To probe the structural consequences of these digenic mutations, we utilized AlphaFold3 to generate a high-confidence model of an AFG3L2-SPG7 heterohexamer (Figure 2A, 2B) and inspected the interactions formed by each wild-type residue of digenic missense mutation pairs. Most of these variants were localized to the matrix-exposed portions of AFG3L2 and SPG7, which contain their ATPase and metalloprotease domains. First, we analyzed the most common digenic pair: *SPG7*:p.(Ala510Val) and *AFG3L2*:p.(Arg702Gln) (Figure 2C). SPG7 A510 is located on a short α-helix (α7) which faces the backside of another α-helix (α6) that forms the SPG7 nucleotide binding site. A510V is predicted to sterically clash with R485 of α6, which could destabilize nucleotide binding. A510 is not located near any inter-subunit contacts, so it is unlikely to have direct effects on oligomerization or assembly. Conversely, AFG3L2 R702 is predicted to be located at an inter-subunit interface between AFG3L2 and SPG7, where it forms polar contacts with SPG7 I671 and AFG3L2 D699. As such, while R702Q is not predicted to sterically clash with its neighbouring SPG7, it could alter these polar interactions to reduce SPG7 binding. The cryo-EM structure of homohexameric AFG3L2 (39) also displays an equivalent R704 in AFG3L2 which mediates similar inter-subunit contacts. As such, *in vitro* work will be useful to address how *AFG3L2* R702Q and *SPG7* A510V might differentially affect AFG3L2-SPG7 assembly compared to AFG3L2-AFG3L2. Two other patients with SPG7 A510V mutations also harbored *AFG3L2* V723M and A659V mutations, respectively. Both residues are located close to the catalytic Zn^2+^ binding site and are not predicted to form inter-subunit contacts. Instead, their mutations may subtly alter AFG3L2 substrate binding and/or protease activity, though neither V723M nor A659V are predicted to introduce severe steric clashes with the AFG3L2 helices that bind Zn^2+^ directly.

**Figure 2.**
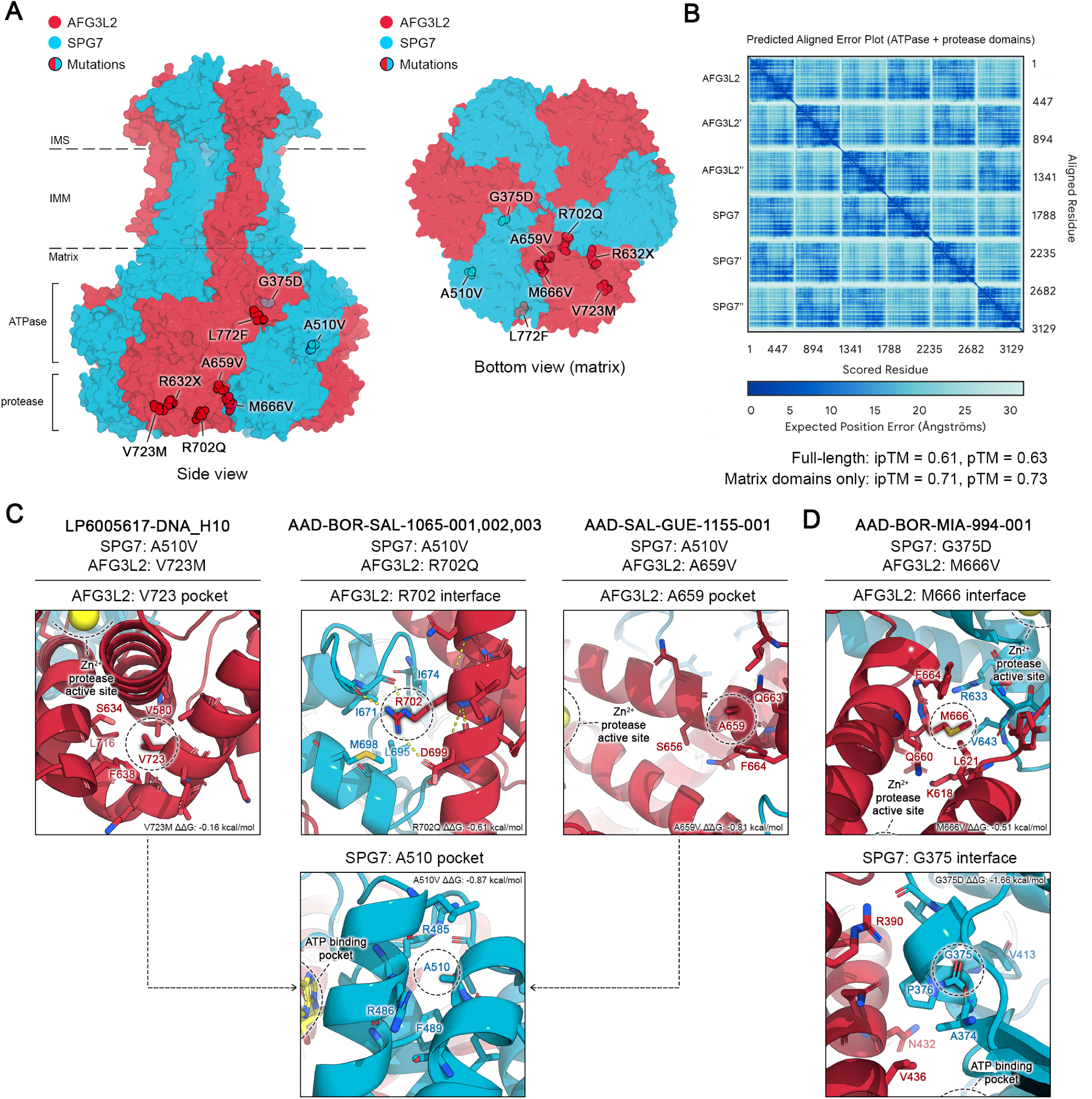
Structural analysis of *AFG3L2* and *SPG7* mutations. **A)** Surface representation of full length AFG3L2-SPG7 heterohexamers as generated by AlphaFold3. Patient mutations were overlaid onto the surface structure and highlighted within the topology of the AFG3L2/SPG7 assembly. Unstructured N-terminal residues with low AlphaFold confidence scores (AFG3L2 a.a. 1-125 and SPG7 a.a. 1-122) were omitted from the surface representation for clarity. All patient mutations were visualized except for SPG7 E484fs, V3delinsVL, and copy number variations. **B)** Predicted aligned error (PAE) plot of the matrix domain AFG3L2/SPG7 assembly from AlphaFold3. Predicted template modeling (pTM) and interface predicted template modeling (ipTM) scores for both full length and matrix complexes were manually annotated. **C)** Structural visualization of selected digenic missense mutations, visualized as atomic interaction sites of the wild-type residue. Residues within 4-5 Å of the target residue were visualized in PyMOL as sticks. Polar contacts formed by Arg702 are depicted as dashed yellow lines. Residues were annotated in Adobe Illustrator. **D)** Structural visualization of SPG7 Gly375 and Met666 interaction sites, as in Figure 1C.

Next, we analyzed another set of digenic missense mutations from one patient: *SPG7* G375D and AFG3L2 M666V, both of which are located at inter-subunit interfaces (Figure 2D). Briefly, SPG7 G375 is located immediately after one β-strand (a.a. 369-374), which is part of the β-sheet that binds nucleotides close to the AFG3L2-SPG7 interface. In our model, SPG7 G375D is predicted to clash with the adjacent SPG7 loop containing V413, which could alter loop dynamics to disrupt inter-subunit AFG3L2-SPG7 contacts and/or nucleotide binding. AFG3L2 M666 is located within the AFG3L2 protease domain, in a hydrophobic pocket that mediates inter-subunit contacts via the Zn^2+^ binding helices of AFG3L2 and SPG7. While M666V is not predicted to directly clash with nearby residues, it could still disrupt the packing of Zn^2+^ binding helices in both inter- and intra-subunit contexts. In support of this model, recombinant mutant M666V AFG3L2 homohexamers displayed reduced proteolytic and ATPase activity compared to wild type AFG3L2 (39). Moreover, recombinant AFG3L2 M666R, a more severely disruptive mutation at the same interface also did not assemble into homohexamers, though the effect of M666V on complex assembly remains unclear. As such, the distinct effects of AFG3L2 M666V on SPG7 binding and proteolysis within the context of the AFG3L2-SPG7 heterohexamer require further studies. Given that the structural mechanisms which govern upstream AFG3L2/SPG7 assembly and folding also remain unclear, *in vitro* studies will be essential to clarify how these digenic mutations interact together within AFG3L2/SPG7 assemblies to affect their assembly, oligomerization, and/or activity.

## Discussion

Our results show that digenic inheritance of potentially pathogenic variants in *SPG7* and *AFG3L2* cause a spectrum of motor neuron and spinocerebellar neurodegenerative disorders. Notably, the majority of *AFG3L2* mutations occur in the peptidase domain indicating that these mutations may abolish proteolytic activity and do not allow the interaction of AFG3L2 with SPG7 subunits into hetero-oligomeric proteolytic complex in the inner mitochondrial membrane. The specific type and position of mutations may explain some of the heterogeneity in the phenotypic spectrum. For instance, the majority of ataxic patients carried *AFG3L2*:p.(Arg702Gln) and *SPG7*:p.(Ala510Val) single nucleotide mutations.

SPG7 performs its function by assembling with its homologous and biological counterpart, AFG3L2, forming a heterohexamer. Both *SPG7* and *AFG3L2* have the highest expression level in the cerebellum and cerebellar hemisphere compared to other regions of the brain (GTEx). Furthermore, they encode highly similar proteins with identical conserved domains. These observations may explain why either homozygous mutations or digenic heterozygous mutations in *SPG7* and *AFG3L2* may lead to the same disorders. Insertions or deletions (indels) in *SPG7* or *AFG3L2* can disrupt the formation and function of the hetero-oligomeric m-AAA protease complex by altering protein structure, stability, or expression levels. Deletions may cause haploinsufficiency, leading to insufficient subunits for complex assembly, while insertions could introduce aberrant sequences, resulting in misfolded or nonfunctional proteins. Such disruptions impair mitochondrial proteostasis, exacerbating dysfunction in digenic inheritance scenarios where pathogenic variants in both genes coexist.

Previous studies have suggested that heterozygous *SPG7* mutations may cause ALS (43, 44). Our findings from one of the largest ALS cohorts analyzed to date, indicate that heterozygous *SPG7* variants alone may not be sufficient to cause ALS. Given our current results of digenic inheritance of *SPG7* and *AFG3L2*, suggest that a likely explanation to these previous reports is that the carriers of heterozygous *SPG7* variants previously reported may also carry variants in *AFG3L2* or in other genes, yet to be discovered. An additional explanation is that undetected structural variants in *SPG7*, which are not often detectable by conventional methods, may have contributed to these cases. Additionally in these previous reports, the heterozygous *SPG7* variants were inherited from asymptomatic parents with age >75 in two of the reported families. Taken together, our results challenge the notion of autosomal dominant inheritance of *SPG7* in ALS.

We previously reported a high number of heterozygous P/LP *SPG7* alleles in HSP patients vs. controls (45). In this case too, the observed overrepresentation of heterozygous *SPG7* variants in patients could be explained by unknown digenic inheritance and other reasons. One additional explanation is that our previous study was based on exome sequencing analysis which is not capable of detecting some classes of disease-causing variants (e.g. e.g. structural variants or variants in intronic regions). For example, CNV analysis of the same data revealed the presence of additional *SPG7* CNVs in two heterozygous carriers of *SPG7* SNVs, showing that they were in fact biallelic carriers of *SPG7* mutations and not heterozygous carriers as we initially reported (25). Furthermore, a deep intronic *SPG7* variant inducing the inclusion of a pseudoexon and premature stop sites was identified in a patient with heterozygous missense *SPG7* variant (46). This variant and other deep intronic variants that may lead to similar effects cannot be detected with exome sequencing. Hence, re-examination and full screening of the entire *SPG7* gene is required for the genetic diagnosis of cerebellopathy patients with at least one detected pathogenic *SPG7* allele. This analysis can then be supplemented by comprehensive analysis for digenic inheritance with *AFG3L2* and other related genes.

Our study has some limitations. We did not perform functional analysis to examine the cellular interaction of SPG7 and AFG3L2 in relevant models, which will be required to fully understand the underlying mechanism. However, previous studies in mice and yeast showed digenic interaction of *spg7-afg3l2* and additive effect of *spg7* and *afg3l2* variants (26, 27), supporting our genetic-clinical findings. Another limitation is that a few variants in our patients, though very rare, are classified as VUS according to ACMG and require pathogenic validation. Additionally, the vast majority of the participants were of European ancestry, indicating a need for studies involving multiple ethnicities. The predominant SCA28 phenotype underscores the complexity of digenic inheritance involving *SPG7* and *AFG3L2* variants, highlighting the need to consider both in genetic counseling, despite the current focus on the SCA28 variant for presymptomatic testing.

## Conclusions

To conclude, our results demonstrating digenic inheritance of *SPG7* and *AFG3L2* in cerebellopathy and corticospinal tractopathy, emphasizing the need to thoroughly study individuals with ataxia and MNDs who carry a single *SPG7* variant. The SPG7-AFG3L2 complex should become a target for translational studies to develop treatments for *SPG7*-associated neurodegenerative disorders.

## Data Availability

All data produced in the present study are available upon reasonable request to the authors

## Abbreviations

HSP: hereditary spastic paraplegia
ALS: amyotrophic lateral sclerosis
SPG7: spastic paraplegia 7
MND: motor neuron disorder
NGS: next-generation sequencing
ACMG: American College of Medical Genetics and Genomics
B: benign
LB: likely benign
VUS: uncertain significance
LP: likely pathogenic
P: pathogenic
SV: structural variant
MLPA: Multiplex ligation-dependent probe amplification
CNV: copy number variations
NDD: neurodegenerative disease

## Funding

This study was funded by CIHR Emerging Team Grant, in collaboration with the Canadian Organization for Rare Disorders (CORD), grant number RN127580–260005, and by a CIHR Foundation grant granted to G.A.R. D.B. is supported by a Humboldt Research Fellowship for Postdocs and the Hertie Network of Excellence in Clinical Neuroscience.

## Acknowledgments

We thank the patients and their families for participating in this study. J.P.R. is supported by Canadian Institutes of Health Research (CIHR) Frederick Banting & Charles Best Doctoral Scholarship (FRN 159279). G.A.R. holds the Wilder Penfield Chair in Neurosciences. Z.G.O. is supported by the Fonds de recherche du Québec–Santé Chercheur-Boursier award and is a Parkinson Canada New Investigator awardee.

## Legends

**Supplementary Table 1 Subjects included in the study**

**Supplementary Table 2 List of potentially pathogenic *SPG7* variants identified in ALS patients and controls.**

**Supplementary Table 3 List of potentially pathogenic *AFG3L2* variants identified in patients and controls.**

**Supplementary Figure 1 Burden testing results based on the ProjectMinE genome sequencing dataset.** The figure derived from Project MinE data browser and shows the exons (orange blocks) in *SPG7* and *AFG3L2* with the variants (triangles) that were observed in the genome sequencing dataset.

## Notes

### Competing Interest Statement

The authors have declared no competing interest.

### Author Declarations

All participants have signed an informed consent form prior to enrollment. The protocol was approved by the McGill University Health Centre (MUHC) Research Ethics Board (REB), a McGill-approved ethics committee based in Montreal, Canada, under project number 2019-4585 and local REB number IRB00010120.

