## Supplementary figures and images for "Digenic inheritance of mutations in *SPG7* and *AFG3L2* causes motor neuron and cerebellar disorders"

### Supplemental Figure 1

#### Slide 1
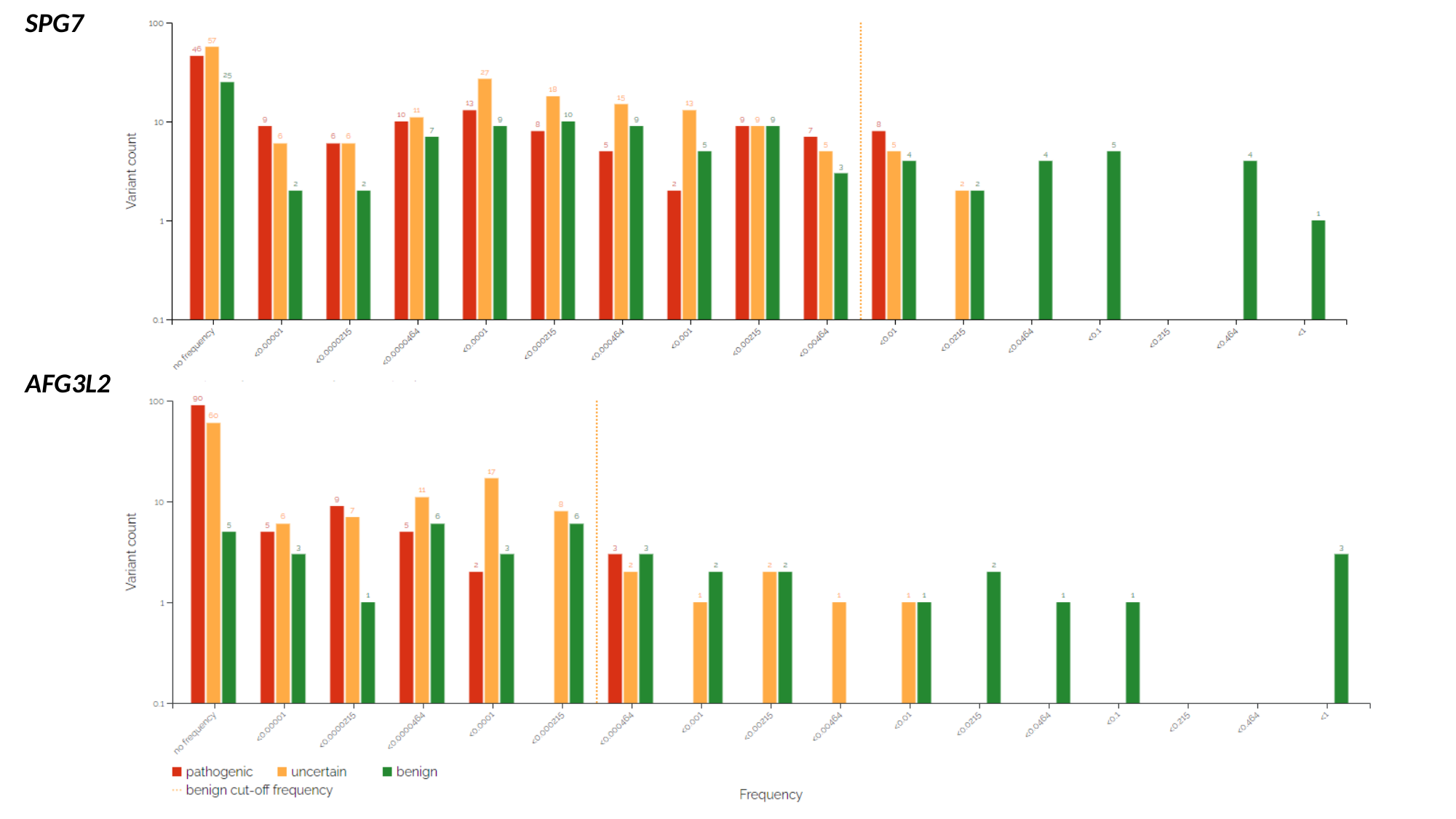

SPG7
AFG3L2
