## Supplemental Table 1 for "Digenic inheritance of mutations in *SPG7* and *AFG3L2* causes motor neuron and cerebellar disorders"

**Supplementary Table 1 Subjects included in the study**

| **Status** | **Cohort** | **Genome Sequencing** | **Exome Sequencing** | **Total** |
| --- | --- | --- | --- | --- |
| Controls | Project MinE + Dr. Rouleau’s lab | 1827 | 580 | 2407 |
| ALS | Project MinE + Dr. Rouleau’s lab | 4356 | 208 | 4564 |
| HSP | CanHSP Cohort | - | 291 | 291 |
| Ataxia | SPATAX cohort, screened in Dr. Durr’s lab | - | 253 | 253 |
| NDD | Dr. Synofzik’s lab | - | 1341 | 1341 |
| NDD | A Cohort, screened in Dr. Synofzik’s lab | - | 5000 | 5000 |
| Rare diseases | GENESIS | - | 12407 | 12407 |

ALS = amyotrophic lateral sclerosis; HSP = hereditary spastic paraplegia; MND = motor neuron disease, NDD = Neurodegenerative disorders
